# An Analysis of Two-Spirit, Lesbian, Gay, Bisexual, Transgender and Queer Research Funded by Canadian Institutes of Health Research

**DOI:** 10.1101/2024.01.09.24301084

**Authors:** Amanda B. Namchuk, Tori N. Stranges, Tallinn F.L. Splinter, Katherine N. Moore, Carmen H. Logie, Liisa A.M. Galea

**Author notes:** **<u>Corresponding Authors</u>** 1., Treliving Family Chair in Women’s Mental Health, Senior Scientist, Centre for Addiction and Mental Health, Professor of Psychiatry, University of Toronto, Toronto, ON, 2., Factor-Inwentash Faculty of Social Work, University of Toronto, Toronto, ON, Canada, United Nations University Institute for Water, Environment and Health, Hamilton, ON, Canada, Centre for Gender & Sexual Health Equity, Vancouver, BC, Canada, Women’s College Research Institute, Women’s College Hospital, Toronto, ON, Canada. indicates co-first authorship (Amanda B. Namchuk, Tori N. Stranges). indicates Co-last authorship (Carmen H. Logie, Liisa A.M. Galea). **Funding:** Funding was awarded to LAMG from the BC Women’s Foundation for this project (AWD-020262). We also gratefully acknowledge funding from the Women’s Health Research Cluster to TNS, ABN and TFLS. CHL acknowledges funding from the Canada Research Chairs program. The authors declare no conflict of interest.

## Abstract

**Purpose:** Gender identity and sexual orientation are essential factors that must be incorporated into health research to ensure we unearth comprehensive and inclusive insights about the healthcare needs and experiences of diverse people. Despite the calls for more focus on sex and gender in health research, scant attention has been paid to gender identity or sexual orientation. Past research found that 0.35% of Canadian Institutes of Health Research (CIHR) grant abstracts mentioned studying lesbian, gay, bisexual, transgender, queer and/or Two-Spirit (2S/LGBTQ+)-specific health outcomes. However, the nature of that research was not explored.

**Methods:** Here we examine the publicly available database of grant abstracts funded by CIHR from 2009-2020 to analyze what type of 2S/LGBTQ+-specific health outcomes would be studied.

**Results:** We found that 58% of awarded grant abstracts mentioned studying sexually transmitted diseases, the majority of which were on human immunodeficiency virus (HIV). Less than 7% of funded 2S/LGBTQ+ grant abstracts mentioned studying cisgender women. Almost 40% mentioned including trans women/girls, and 30% mentioned including trans men/ boys. None of the studies examined mentioned work with the Two-Spirit community.

**Conclusion:** These results reflect larger social and health inequities that require structural level changes in research to support lesbian, bisexual and queer women’s health.

## Introduction

Two-Spirited, Lesbian, Gay, Bisexual, Transgender, Queer or Questioning (2S/LGBTQ+) is an umbrella term that includes individuals who have both diverse sexual orientations as well as gender identities. Gender identity and sexual orientation are critical considerations for health research, especially for the 2S/LGBTQ+ community^1^. Gender plays a role in healthcare seeking practices, symptom perception by clinicians, disease diagnosis, and treatment^2–4^. However, the ways in which diverse gender identities interact with sex hormones to shape one’s physiology is not as well known. Outside of a healthcare setting, a mismatch between gender identity, expression and societal gender roles can itself be a stressor^5^, which may contribute to differences in stress reactivity between 2S/LGBTQ+ individuals and their heterosexual counterparts^6^. Further, research on allostatic load demonstrates the energetic burden put on physiological systems when environmental stress exceeds an individual’s ability to cope. This burden is particularly apparent for the 2S/LGBTQ+ community who live in environments of heterosexism, cissexism, and transphobia^6^. Stress hormones play a pivotal role in this process. For some transgender individuals who experience a distressing mismatch between their gender and sex assigned at birth, gender-affirming hormone therapy (GAHT) can help to relieve the psychological burden, yet little is known about how GAHT affects other domains such as cognition, cardiovascular health, and skeletal health^7–9^. Indeed, surveys conducted during the initial year of the global COVID-19 pandemic, demonstrated a larger burden on the 2S/LGBTQ+ community on anxiety, perceived stress, depression and loneliness scores^10^. Collectively, understanding the unique needs of diverse gender identities and sexual orientations in the 2S/LGBTQ+ community is imperative in addressing the healthcare needs of this population.

Further, stigma and discrimination toward persons with diverse sexual orientations are stressors and social determinants of health that result in poorer health outcomes compared to heterosexual counterparts both by producing barriers to accessing health and social services and through the experience of stigma itself^11–14^. Although 2S/LGBTQ+-affirming healthcare improves health outcomes and mitigates harm caused by stigma and discrimination, 2S/LGBTQ+□community members continue to experience increased disease risk and poorer health outcomes as a result^15,16^. Experiences in healthcare are intersectional, as the 2S/LGBTQ□+□community’s reduced access to power, resources and opportunities are further compounded with other marginalized identities, such as race and class^17–19^. Indeed, gender inequality across societies is linked to increased mental health indices, and result in cortical thickness differences in limbic regions between women and men^20^, but to our knowledge these data have not extended to other genders or sexual orientation.

Given the importance of studying 2S/LGBTQ+ health questions, it is concerning that recent research from our group revealed that less than 1% of all the Canadian Institutes of Health Research (CIHR) Operating and Project Grants were awarded to projects considering 2S/LGBTQ+ health from 2009-2020^21^. This finding highlights that 2S/LGBTQ+□health requires explicit attention and funding dollars to address the long-standing social and health disparities within these populations.

However, not all 2S/LGBTQ+ persons are included equally in research. For instance, Browne and Nash^22^ highlight that the methods through which statistical data are gathered on sexuality (e.g., mainly through market research or census data on couples) can result in a biased sample of predominantly white, affluent, middle class, gay men who live in specific urban areas. Further, Coulter et al^23^ reported that the lack of funded research (particularly by the United States National Institutes of Health Research [NIH]) on and about 2S/LGBTQ+ health contributes to the perpetuation of health inequities. Poteat et al.^24^ note that most data on 2S/LGBTQ+ health focuses on human immunodeficiency virus (HIV) among gay, bisexual and men who have sex with men (gbMSM). As a result of this focus, there are gaps in knowledge for gbMSM health besides HIV research, and large gaps for lesbian, bisexual, and queer (LBQ) women across the health spectrum^24^. These findings are consistent with evidence that there is nine times more health research published in males compared to that published in females in both human and animal research^25^. To address knowledge gaps regarding 2S/LGBTQ+ funding patterns in Canada, we searched the publicly available database of grant abstracts funded by CIHR to analyze what 2S/LGBTQ+ populations and specific health outcomes are mentioned in the abstracts from 2009 to 2020. The goal of this analysis was to determine, within CIHR-funded 2S/LGBTQ+ research abstracts, what populations and health issues were mentioned.

## Methods

In a previous study, we examined the published public abstracts of all Operating and Project Grants funded by CIHR from 2009 to 2020 using the CIHR Funding Decisions Database (see Stranges et al., 2023 for detailed methodology). Operating and Project Grants are CIHR’s major funding competitions (akin to R01 mechanisms through the NIH). Using the abstracts previously identified as mentioning a health issue relevant to the 2S/LGBTQ+ community or using 2S/LGBTQ+ participants (n=33), we manually coded grant abstracts into additional categories based on the content of the abstract. We coded relevant abstracts for participant gender (gender identity of participants was outlined), participant sexual orientation (sexual orientation of participant was outlined) and the proposed topic to be studied (e.g., HIV, health care barriers, mental health). Grant abstracts that did not clearly fall into one or more categories (n=4) were discussed between the three lead coding team members and were coded appropriately once a unanimous decision was met. Grant abstracts that proposed studying an issue relevant to the 2S/LGBTQ+ community but did not study the community members directly were excluded from this analysis (n=2). Additionally, one computer modeling study was included in the topic analysis but not the participant analysis, as it did not include human subjects. Therefore, 30 abstracts were included in the participant gender and sexual orientation analyses and 31 abstracts were included in the topic analysis.

Grant abstracts were coded for mentioning including cisgender women/girls, cisgender men/boys, transgender (trans) women/girls, trans men/boys, and/or nonbinary individuals. Abstracts that mentioned including participants of multiple genders were coded for each gender mentioned. In the case that an abstract mentioned including women/girls or men/boys but did not specify whether the participants would be trans- or cisgender, the participants were assumed to be cisgender. Abstracts were additionally coded for mentioning including gbMSM only or additional sexual orientations beyond gbMSM. All abstracts that included multiple sexual orientations were coded as including additional sexual orientations.

For the topic analysis, abstracts were allocated into 7 research topics that included: 1) HIV (any grant that considered HIV or preexposure prophylaxis [PrEP] for HIV prevention); 2) other infectious diseases, not including HIV (e.g., HPV, Hepatitis C); 3) accessibility and medical experiences (e.g., health care accessibility and experiences accessing care); 4) risk and resilience (e.g., socio-cultural and environmental factors that influence health); 5) mental health and substance use; 6) brain development (e.g., how hormone therapy impacts brain development); and 7) gbMSM blood pathogen profiles (e.g., pathogens transmitted through blood donation).

## Results

### Participant Gender and Sexual Orientation

First, we categorized the gender and sexual orientation of the participants to be recruited for the proposed research. Participant gender was mentioned in three-quarters (76.7%) of abstracts (Fig. 1A) and sexual orientation was mentioned in 60.0% of abstracts (Fig. 1D).

**Figure 1.**
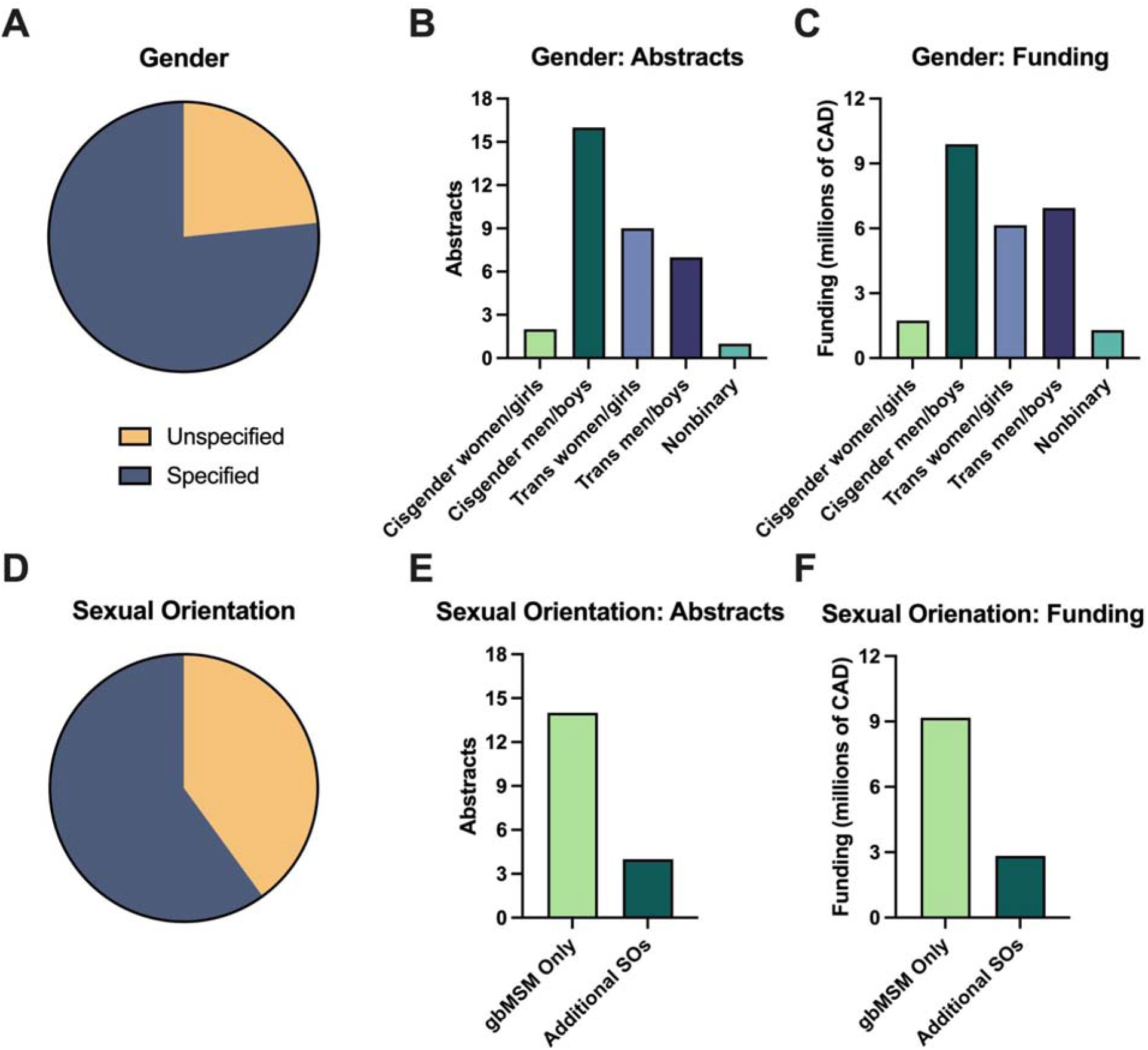
Gender (A-C) and sexual orientation (D-F) of participants to be recruited for studies proposed in grant abstracts. N=30 and total funding was 20.1 million CAD. gbMSM only: gay, bisexual, and other men who have sex with men. Additional SOs: additional sexual orientations (SOs) beyond gbMSM. Abstracts that mentioned including participants of multiple genders were coded multiple times. Abstracts that mentioned including participants of multiple sexual orientations were coded as including additional SOs.

No abstracts mentioned including Two-Spirit individuals (although one abstract proposed research in Indigenous communities specifically).

Of the grant abstracts that specified a participant gender, study participants included 69.6% cisgender men/boys (n=16), 39.1% trans women/girls (n=9), 30.4% trans men/boys (n=7), 8.7% cisgender women/girls (n=2), and 4.3% nonbinary persons (n=1; Figure 1B). Of abstracts that specified participant sexual orientation, 77.8% (n=14) reported research participants that were exclusively gbMSM and 22.2% of studies (n=4) mentioned including research participants of additional sexual orientations within the 2S/LGBTQ+ community beyond gbMSM, including lesbian, bisexual, and queer (LBQ) individuals (see Figure 1E).

### Funding levels

We next examined the funding levels for these abstracts by category. Grants that mentioned studying cisgender men/boys within the 2S/LGBTQ+ community received 53.3% of 2S/LGBTQ+ funding (9.9 million CAD). In contrast, 6.7% of 2S/LGBTQ+ funding (1.7 million CAD) mentioned including cisgender women/girls. Grants that mentioned studying trans women/girls received 30.6% (6.2 million CAD) of funding and grants that mentioned studying trans men/boys received 23.3% (6.9 million CAD) of funding, respectively. Abstracts that mentioned nonbinary participants received 6.5% (1.3 million CAD) of funding.

### Research Topics

Grant proposal abstracts covered a variety of research topics including: HIV; other infectious diseases; accessibility and medical experiences; mechanisms of disease risk and resilience; mental health and substance use; brain development; and gbMSM blood pathogen profiles (Fig. 2).

**Figure 2.**
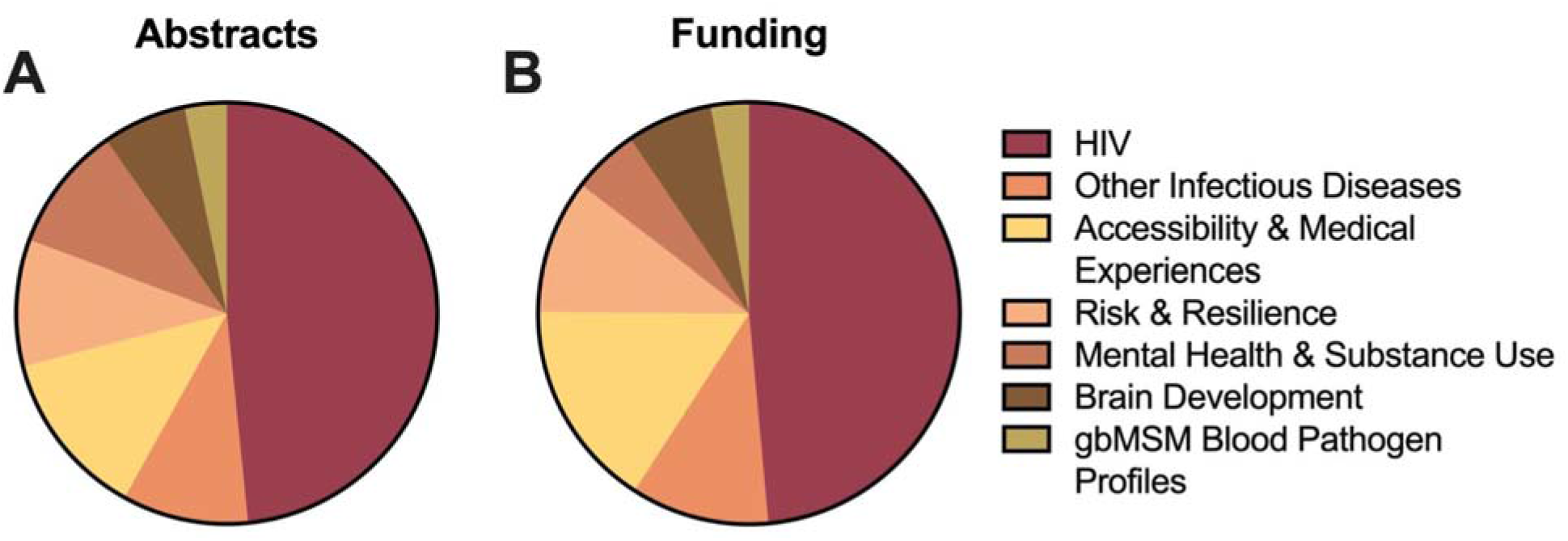
Distribution of proposed topics of study mentioned in 2S/LGBTQ+ abstracts funded by CIHR in A) abstracts and B) funding dollars. N=31 and total funding was 20.1 million CAD. HIV: human immunodeficiency virus. gbMSM: gay and bisexual men who have sex with men.

Among funded CIHR research abstracts, the largest health issue mentioned was HIV, which was a focal area in 48.4% (n=15) of studies. HIV was by far the most commonly funded research topic with a 9.9 million CAD (48.5%) investment, comprising nearly half the funding dollars awarded (Fig. 2). An additional 9.7% of abstracts mentioned examining infectious diseases other than HIV (n=3; 2 on Human Papillomavirus [HPV] and 1 on Hepatitis C) and received 10.6% of funding dollars (2.2 million CAD). When combined, a total of 58.1% of abstracts mentioned infectious diseases (n=18).This accounts for 59.1% (12.1 million CAD) of total funding awarded (Fig. 2).

The remaining 41.9% of grant abstracts addressed other topics (Fig. 2). The most common research topic after HIV and infectious diseases was accessibility and medical experiences, comprising 12.9% of abstracts (n=4) and receiving 16% of funding dollars (3.3 million CAD). One-tenth (9.7%) of the total abstracts examined mentioned disease risk and resilience (n=3) and another 9.7% mentioned mental health and substance use (n=3). Proposals on these topics received 10.3% (2.1 million CAD) and 5.1% of funding dollars, respectively. Grants mentioning brain development (n=2) comprised 6.5% of abstracts and received 6.6% of funding dollars (1.3 million CAD). Finally, 3.2% of abstracts mentioned gbMSM blood pathogen profiles (n=1). This proposal received 2.9% of funding (0.6 million CAD) (Fig. 2).

### Other Noteworthy Findings

Of the 15 grant proposal abstracts that mentioned studying HIV, only 3 (20.0%) mentioned that they included participants other than cisgender men, while 2 grant abstracts mentioned including cisgender women. Of the 3 additional abstracts on non-HIV infectious diseases, the 2 investigating HPV included cisgender men only (gbMSM) and the 1 investigating Hepatitis C did not specify participant gender or sexual orientation.

Of the 14 proposals examining gbMSM only, 13 (92.9%) mentioned examining HIV or HPV, both sexually transmitted infections (STIs). Both proposals investigating the impact of sex hormones on brain development included adolescent trans boys and girls. Of the 3 abstracts that mentioned studying mental health or substance use, none specified participant gender and only 1 (33.3%) specified participant sexual orientation.

## Discussion

Overall, our analysis of CIHR-funded abstracts that mentioned 2S/LGBTQ+-specific health outcomes revealed that the majority of the awarded grant abstracts on 2S/LGBTQ+ health addressed STIs, and the majority of those mentioned studying HIV. Additionally, STI proposed research was primarily to occur in cisgender men. Health issues, other than STIs, are overlooked among sexual and gender diverse populations in Canada and other global regions^24^ (Poteat et al., 2021). It is critical to move beyond a focus on HIV when considering 2S/LGBTQ+ health research. For instance, there have been calls to focus on a life course perspective and address non-communicable diseases^24^. Other areas of urgent concern include healthy 2S/LGBTQ+ aging, health of 2S/LGBTQ+ incarcerated persons, sexual pleasure, and wellbeing (including but expanding beyond HIV and STIs), and structural determinants of 2S/LGBTQ+ health (e.g., social norms, healthcare provider competence). Our study findings are important as they highlight that within 2S/LGBTQ+ research, the majority is centered on studying STIs, and clearly broader research categories are needed.

Understanding the intricate relationship between diverse gender identities and sex hormones is a complex field that warrants further exploration. The intersection of gender identity, expression, and societal gender norms can become a significant stressor^5^. This stress may contribute to variations in stress reactivity between individuals within the 2S/LGBTQ+ community and their heterosexual counterparts. Further, the burden of allostatic load is particularly pronounced for the 2S/LGBTQ+ community^6^. Stress hormones play a pivotal role in this complex process, acting as mediators between environmental stressors and the body’s physiological response. For transgender, Two-Spirit, and non-binary community members, GAHT must also be considered; however, GAHT continues to be underfunded and under researched. We found only 2 proposals investigating the impact of sex hormones on brain development across over a decade of CIHR funding. Despite its proven psychological benefits, there remains a dearth of knowledge concerning how GAHT impacts other domains of health, including cognition, cardiovascular health, and skeletal health. As our understanding of gender-affirming interventions continues to evolve, research in these areas becomes essential to provide comprehensive care and support. This will contribute to a more holistic understanding of the intersection between gender identity, hormone therapies, and overall well-being.

The present data also signals health research inequities at the intersection of gender identity and sexual orientation. Lesbian, bisexual and queer (LBQ) women are severely underrepresented in CIHR funded health research, as they were mentioned in less than 7% of grant abstracts. This marginalization of women in health research, including specifically LBQ women, is not a new phenomenon. These findings are consistent with the broad underrepresentation of all women in clinical trials and in published research^25–27^ and LBQ women in health research specifically^24^. Similar analyses in the US and in the European Union have indicated underfunding of female health^28^ and have led to calls for a more concerted effort to fund women’s health research^29^. The low inclusion of LBQ women has also been noted in research with women living with HIV^30^, in sexual health research and HIV research^31^. The omission of LBQ women from research is a form of structural intersectional stigma that itself re/produces health inequities. Health equity is not possible when LBQ women are left out of the research conversation.

## Conclusion

It is challenging to understand why one-quarter of studies did not report gender and 40% did not include sexual orientation in this era of heightened sex and gender based analysis (SGBA) focus at CIHR. SGBA has been mandated since 2019 for CIHR Project grants and was introduced to the Canadian academic community in 2010. Requiring accurate reporting of sexual orientation, gender identity and sex characteristics (SOGISC) is key for reliable and valid research, and there is now plentiful guidance and resources provided for researchers to undertake ethical, strengths-focused and community-engaged research with 2S/LGBTQ+ persons to ensure that research advances social justice and health equity (Logie, van der Merwe, & Scheim et al., 2022). It is important to acknowledge that categories within gender identity are rapidly evolving and that many questionnaires and databases that include this information on gender identity and sexual orientation are dated (Bauer et al., 2017). However, it is equally important to understand that the gender and health issues bias in what is funded in this data mirrors larger social and health inequities based on gender inequity and stigma. To change outcomes will require structural level changes in research support for LBQ women’s health and Two-Spirit support. This shift must include research led by and for LBQ women and additional support for non-HIV related research with gbMSM, Two-Spirit, trans and non binary communities. The exclusion of Two-Spirit people from health research continues to perpetuate a concerning gap in culturally competent and inclusive health care practices and policies. What is funded by a national funding body reflects society’s values and priorities, and studies that make visible what (and who) are not funded can help the research community to recalibrate and support understudied 2S/LGBTQ+ topics and communities.

## Data Availability

All data produced are available online at https://webapps.cihr-irsc.gc.ca/decisions/p/main.html?lang=en#sort=namesort%20asc&start=0&rows=20

